# Detection of oncometabolites 1-methylnicotinamide, nicotine imine and N-Methylnicotinium in nails of oral cancer patients and prediction of them as modulators of DNMT1

**DOI:** 10.1101/2020.09.20.20198101

**Authors:** Anwesha Deep Dutta, Ajay Kumar, Kiran Lokhande, Manmohan Mitruka, Jayanta K. Pal, Sachin C. Sarode, Nilesh Kumar Sharma

## Abstract

**Background:** The prominent among various existing views on the role of nicotine and nicotine-metabolized products in Oral squamous cell carcinoma (OSSC) is metabolic adaptation that allows the use of methyl-donor S-adenosylmethionine (SAM) for non-epigenetic purpose including the methylation of nicotinamide and nicotine. In fact, channeling of SAM for generation of 1-methylnicotinamide (1-MNA) and methylated nicotine products is seen as a key event in cancer cells that allows favorable epigenetic states by forcing DNA hypomethylation. A better perception of such events can be appreciated by analyzing samples like nail, which represents a perfect biological material for studying long-term metabolic reflections of the body.

**Methods:** Potential nicotine-metabolized products and 1-MNA in nails of OSCC patients were analyzed by using a novel approach of Vertical tube gel electrophoresis (VTGE)- assisted purification followed by their identification by LC-HRMS. Further, these identified nicotine metabolized products and 1-MNA were evaluated for their molecular interactions with known methyltransferases including cytosolic nicotinamide methyltransferase (NNMT), DNA methyltransferase (DNMT)1 and histone methyltransferases by molecular docking and molecular dynamics simulation (MDS) analyses.

**Results:** Our data suggests the presence of N-methylnicotinium ion and nicotine imine in the nail samples of OSCC patients. Further, 1-MNA is also detected in the nails as a major enzymatic product of a known detoxifying enzyme NNMT. Molecular docking of all nicotine and nicotine metabolized products with DNMT1revealed a specific binding affinity of nicotine imine only with a -6.2 Kcal/Mol docking energy. Importantly, binding of nicotine imine is within the CXCC regulatory domain of DNMT1 and it displays molecular interactions with the key amino acid residues, namely ARG690, PRO574, VAL658, PRO692 and ALA695. Furthermore, MDS data corroborated well with the specific binding affinity of nicotine imine to DNMT1 obtained by docking analysis.

**Conclusion:** Identification of N-methylnicotinium ion, nicotine imine and 1-MNA in nail samples indicates their potential as predictive and detectable biomarkers for OSCC. Molecular docking and MDS data lead us to propose a role of nicotine imine in modulating the activity of DNMT1. These data further suggest a novel understanding on the role of nicotine metabolized products in modulating DNA methylation that may contribute to tumorigenicity in oral cancer patients.

## INTRODUCTION

According to a global cancer report, OSCC is a global health problem and in 2018, the total new cases of lip and oral cavity cancer and new deaths were 3,54,864 and 1,77,384, respectively (Bray et al., 2018). Thus, OSCC is regarded as a global health problem demanding imperative attention of multi-disciplinary nature (Nakajima, 2007; Benowitz et al., 2009; Chang et al., 2017; Sarode et al., 2018; Jiang et al., 2019; Miranda-Filho et al., 2020; Sarode et al., 2020). OSCC is deemed as a multi-factorial disease with a predominant association with tobacco consumption. Nicotine is a major and addictive constituent of tobacco and is reported to be metabolized into components including nicotine imine, N-methylnicotinium ion and cotinine by cytochrome P450 enzymes and methyltransferases by hepatic and epithelial cells (Nwosu et al., 1988; Berkman et al., 1995; von Weymarn et al., 2002; Kilgore et al., 2013; Jeltsch et al., 2016; Miletić et al., 2017; Yu et al., 2019). Evidences suggest that nicotine as such is not a carcinogen but upon metabolism, its products contribute to metabolic-epigenetic modifications relevant to malignant transformation and progression (Nwosu et al., 1988; Berkman et al., 1995; von Weymarn et al., 2002; Kilgore et al., 2013; Jeltsch et al., 2016; Miletić et al., 2017; Yu et al., 2019).

Among cancer and normal cells, an axis that links metabolic and epigenetic regulation is known to exist to promote growth and proliferation (DeBerardinis et al., 2008; Hsu and Sabatini, 2008; Hanahan and Weinberg, 2011; Ulanovskaya et al., 2013; Feinberg et al., 2016; Yu et al., 2018; Kim et al., 2019). In fact, metabolic reprogramming is different in cancer cells that lead to pro-tumor intracellular landscape. Among various metabolic adaptations in cancer cells, involvement of various classes of methyltransferases including cytoplasmic nicotine methyltransferase (NNMT) and nuclear methyltransferases that include DNA methyltransferase and histone methyltransferase is imperative (Feinberg and Vogelstein, 1983; Christmane et al.., 2002; Lopez-Serra and Esteller, 2008; Palii et al., 2008; Wu et al., 2008; Ulanovskaya et al., 2013; Kraus et al., 2014; Cui et al., 2020; Gissi et al., 2020). There are limited attempts that show the connection between metabolic and epigenetic landscape including the use of SAM to drive the need of NNMT enzyme that leads to the less availability of SAM for methylation activity of DNMT1 upon the DNA (Wu et al., 2008; Ulanovskaya et al., 2013; Kraus et al., 2014; Cui et al., 2020). Data support that inhibition of DNMT1 is linked with the global hypomethylation and associated with the pro-tumor transcriptional state in cancer cells (Feinberg and Vogelstein, 1983; Christmane et al.., 2002; Lopez-Serra and Esteller, 2008; Palii et al., 2008; Gissi et al., 2020). In spite of knowledge on the existence of intracellular signaling axis that links the use of SAM, activation of NNMT and DNA hypomethylation, molecular and clinical relevance of NNMT-induced abundance of oncometabolite 1-MNA in oral cancer is completely lacking.

Nail has been considered as the best material for bio-monitoring of the pathophysiological changes over a period of time. Nail clipping samples reflect the biochemical changes over 12 to 18 months period of time, and therefore it is most commonly used for detection of prolonged exposure of external noxious agents to the body (Oyoo-Okoth et al., 2010; Krumbiegel et al., 2016; Mitruka et al., 2020; Sharma et al., 2020). Interestingly, S-nicotine metabolized products including nicotine imine and N-methylnicotinium ion are reported to some extent in different tissues and biological fluids (Nwosu et al., 1988; Berkman et al., 1995; von Weymarn et al., 2002; Kilgore et al., 2013; Jeltsch et al., 2016; Miletić et al., 2017; Yu et al., 2019). Nonetheless, these nicotine products nicotine imines and N-methylnicotinium are not reported in conventional nail materials of OSCC and their biological relevance is not discussed. Furthermore, our understanding on the role of these oncometabolites including nicotine imine and N-methylnicotinium is completely missing and further investigation may establish their role in changing the metabolic-epigenetic landscape in favor of OSCC. Therefore, relevance of determining the role of oncometabolites 1-MNA, nicotine imine and N-methylnicotinium in modulating the activities of NNMT and DNMT1 in the background of favorable metabolic reprogramming is highly warranted by employing in silico and in vitro approaches.

Additionally, evidence on the detection of nicotine metabolized products in nails and other biological fluids are limited due to the non-availability of appropriate purification and detection approaches. Based on above understanding and limitations, we aim to identify nicotine metabolism products and 1-MNA in nails of OSCC and their molecular interaction with different cytosolic and nuclear methyltransferases including NNMT and DNMT1 to understand-epigenetic states leading to metabolic adaptations during oncogenesis in OSCC.

## MATERALS AND METHODS

### Study Population

OSCC patients (n=5) and healthy subjects (n=6) were recruited from Dr. D. Y. Patil Dental College and Hospital, Pune, India. Institutional Ethics Committee of Dr. D. Y. Patil Vidyapeeth, Pune (Ref.NoDYPV/EEEEEC/245/2019) approval was obtained before commencement of the study. All the participating study population was appraised of the objectives of the study and informed consent was obtained prior to investigations.

### Purification and identification of nail metabolites

The primary goals were to study the nail lysate in the context of nicotine and nicotine-derived metabolites with the assistance of a novel vertical tube gel electrophoresis (VTGE) metabolite purification system and identification of these metabolites with the help of LC-HRMS (Sharma et al., 2019; Mitruka et al., 2020). A working model of novel VTGE system is presented in Figure S1. In brief, fingernail clippings were lysed in 800 µl of extraction buffer, 20mM Tris-HCl (pH-8.5) containing 2.6M Thiourea, 5M Urea and 800 µl Beta-Mercaptoethanol. The prepared nail lysates were purified by using VTGE metabolite purification system. In brief, RPC18 column (Zorbax, 2.1 × 50 mm, 1.8 µm) was used for liquid chromatography (LC) component. Furthermore, nail metabolites were submitted to positive electrospray ionization (ESI) M-H mode by mass spectrometer component as MS Q-TOF Quadrupole time-of-flight mass spectrometry (Q-TOF-MS).

### Molecular Docking Study on Oncometabolite Biomarker and Proteins

To perform molecular interaction studies, oncometabolites and known inhibitor 1-Methylnicotinamide (PUBCHEM CID: 457), Nicotine imine (PUBCHEM CID: 431), N-Methylnicotinium ion (PUBCHEM CID: 430), 5’-Aza-2’-deoxycytidine (PUBCHEM CID: 22841634) were retrieved as ligand for molecular docking. The ChEBI (https://www.ebi.ac.uk/chebi/) database was used to download the structure of ligands in SDF format. Then, the retrieved compounds were converted into PDB file format with 3-dimentional coordinates using OpenBable software. The targeted structures of potential methylatranferases including NNMT Enzyme (PDB ID 3ROD), Human Euchromatic Histone Methyltransferase 2 (PDB ID:2O8J), Methyltransferase domain of human euchromatic histone methyltransferase 1 (PDB ID:4I51), a non-set domain nucleosomal histone methyltransferase (PDB ID 1NW3), Human euchromatic histone methyltransferase I SET Domain (PDB ID 2IGQ), Human DNMT1 (PDB ID4WXX) were taken from Protein Data Bank (https://www.rcsb.org) with 3.05 Å resolution. All the Hetatoms were removed prior to docking. The selected protein receptors were subjected to protein preparation by using Autodock 4.2 software (Morris et al., 2009). This included removal of water molecules, bond correction, assigning AD4 type atoms, addition of polar charges and addition of Kollman charges. For in silico studies, AutoDock Vina software was used to perform molecular docking of the ligands with the targeted protein (Trott and Olson, 2010). AutoDock Vina comes with the feature of the calculation of grid maps automatically. The binding cavity of the receptor was first determined using blind docking which involves covering the entire receptor with the grid box of appropriate size. The docking procedure involved the systemic conformational expansion of the ligand followed by the placement in the receptor active site. The best conformation was determined by the binding affinity and interaction patterns of the complex. After successful docking, the ligand-receptor complex was viewed in Discovery Studio viewer (DSV3, 2010).

### Molecular dynamics Simulation

The molecular simulation dynamics was carried for the complex of Nicotine imine (PubChem CID: 431) and DNMT1 enzyme (PDB ID 4WXX), using Desmond software to assess the stability of ligand and protein complex. We performed 10ns MD simulation to see the base conformational changes in the binding pocket of Nicotine imine: NNMT complex. Using a system builder of Desmond in the Maestro program, the system for all the complexes was immersed in a water-filled orthorhombic box of 10 Å spacing. The complex system of Nicotine imine-DNMT1 enzyme had water molecules of 8944 using an extended three-point water model (TIP3P) with periodic boundary conditions. The total charge of the solvent system was neutralized by adding 2 sodium (Na+) of concentration 4.066 mM. These studies were carried out with a run of 10ns and temperature 300K, considering certain parameters including integrator as MD (Schrödinger, 2019). The conformational changes upon binding of Nicotine imine with DNMT1 enzyme were recorded by using the 1000 trajectories frames generated during the 10ns MD simulation (Schrödinger, 2019).

## STATISTICAL ANALYSIS

Data are presented as the mean ± SD of at least three independent experiments. Differences are considered statistically significant at P < 0.05, using a Student’s t-test.

## RESULTS

### LC-HRMS analysis and detection of nicotine metabolized products

In literature, limited data is available that indicate the role of nicotine and nicotine metabolized products in promoting the cancer metabolic shift that may help in the tumorigenesis. Furthermore, metabolite profiling in view of SAM as a methyl-donor and generation of oncometabolites such as 1-MNA and methylated nicotine product is not reported in nails and other biological fluids in OSCC. Therefore, the use of VTGE, a novel metabolite assisting tools discovered in our lab helped to purify the nail metabolites and subsequently subjected to LC-HRMS. The positive ESI extracted ion chromatogram (EIC) ion spectra is presented in Figure 1 A, B, and C for N-methylnicotinamide, Nicotine imine and N-methylnicotinium ion, respectively. Here, specific fragment ion spectra for N-methylnicotinamide (118.05255, 136.06311), nicotine imine (307.0689) and N-methylnicotinium ion (344.1329) are detected and validated during the mass ionization. It is important to mention that as such nicotine is not detected during the metabolite profiling of nail samples. In supplementary data, positive ESI mode water loss adduct TIC of VTGE purified nail metabolites is given for healthy subjects (Figure S2 A) and OSCC (Figure S2 B). In this TIC, oncometabolite N-Methylnicotinamide is detected. Furthermore, positive ESI mode sodium adduct TIC of VTGE purified nail metabolites is given for healthy subjects (Figure S3 A) and OSCC (Figure S3 B). In this TIC, nicotine metabolized products nicotine imine and N-methylnicotinium ion are noticed.

**Figure 1.**
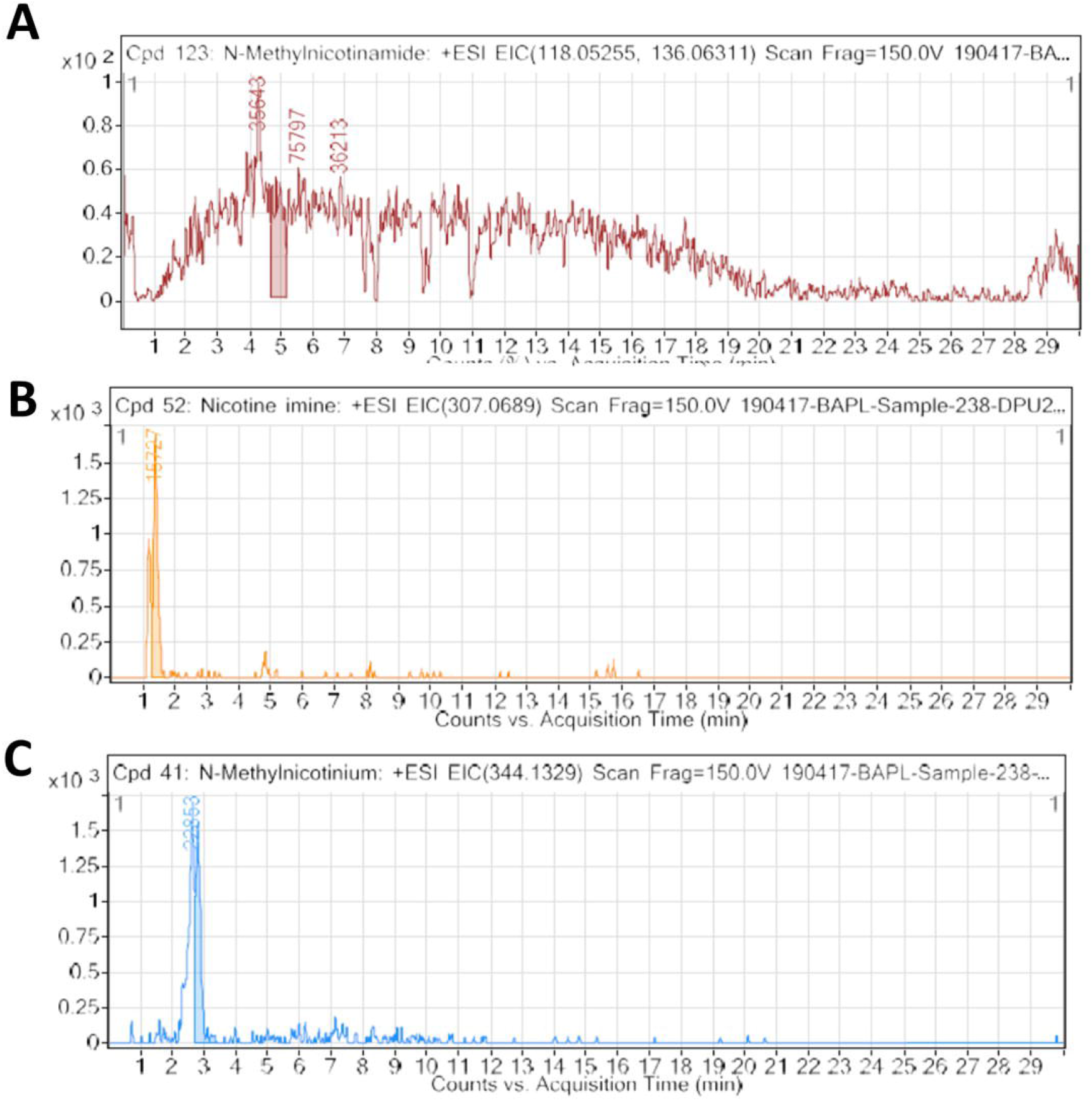
Oncometabolites 1-Methyl nicotinamide, Nicotine imine and N-Methylnicotinium ion are detected in nails of OSCC patients. The above figure shows the Extracted Ion Chromatograph (EIC) from the LCMS of metabolites obtained from the nail clippings of OSCC Patients. **(A)** 1-Methyl nicotinamide. **(B)** Nicotine imine. **(C)** N-Methylnicotinium ion. These oncometabolites are in abundance in nail lysates purified by VTGE methodology from OSCC patients. Here, ESI MS spectra are collected in + ESI mode.

### Molecular docking of N-methylnicotinamide, Nicotine imine and N-methylnicotinium ion with cytosolic and nuclear methyltransferases

Of late, various classes of cytosolic and nuclear methyltransferases have been implicated in detoxification (NNMT enzyme) and DNA methylation (DNMT1). Interestingly, a molecular link between oncometabolites such as N-methylnicotinamide, Nicotine imine and N-methylnicotinium ion and these potential methyltransferases is completely missing. Hence, AutoDock Vina based molecular docking of these oncometabolites such as N-methylnicotinamide (Table 1), Nicotine imine (Table 2) and N-methylnicotinium ion (Table 3) was performed upon NNMT Enzyme (PDB ID: 3ROD), human Euchromatic Histone Methyltransferase 2 (PDB ID: 2O8J), methyltransferase domain of human euchromatic histone methyltransferase 1 (PDB ID: 4I51), catalytic domain of human DOT1L, a non-set domain nucleosomal histone methyltransferase (PDB ID: 1NW3), human euchromatic histone methyltransferase I SET Domain (PDB ID: 2IGQ) and Human DNMT1 (PDB ID: 4WXX). Our results indicated that these oncometabolites Nicotine imine (binding energy -6.2 Kcal/Mol), N-methylnicotinium ion (binding energy -5.4 Kcal/Mol), and N-methylnicotinamide (binding energy 5.6 Kcal/Mol) showed specific and functional molecular interactions with only DNMT1 enzyme that is known to perform the DNA methylation and in turn involved in cancer epigenetic state. Importantly, we performed docking of a known DNMT1 inhibitor 5’-Aza-2’-deoxycytidine (PUBCHEM CID: 22841634) and almost similar binding energy -7.0 Kcal/Mol) compared to nicotine imine was recorded. This molecular docking data encouraged to understand the interacting amino acid residues in case of the tested oncometabolites and known DNMT1 inhibitor 5’-Aza-2’-deoxycytidine. We used Discovery studio visualizer to estimate the nature of polar bonds, number of polar bonds, bond distance and involved amino acid residues at the molecular docking position between oncometabolites and DNMT1. Precisely, nicotine imine (Figure 2) and 1-MNA (Figure 3) showed specific molecular interactions at the regulatory site of DNMT1 with specific residues, namely ARG690, PRO374, GLU566, PRO692, VAL658 and ALA695. With reference to N-methylnicotinium ion, a completely different molecular interaction with DNMT1 was observed in comparison to nicotine imine and 1-MNA by showing specific binding to amino acid residues namely, GLU1168, PHE-1145, PRO1225 and GLU-1266 (Figure 4). In essence, these interacting amino acid residues are known to occupy within the catalytic domain of DNMT1.

**Table 1:**
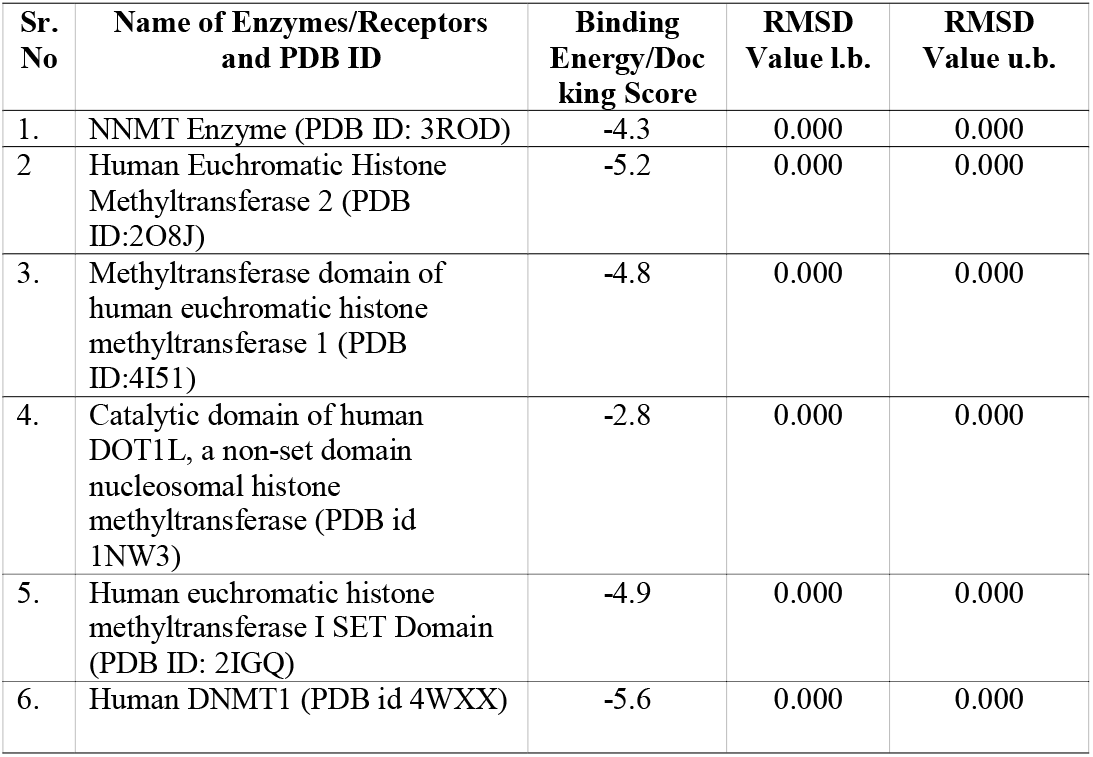
Details on molecular docking by AutoDock Vina of 1-Methylnicotinamide upon selected methyltransferases enzymes involved in epigenetic-metabolic axis that support cancer growth and proliferation. 1-Methylnicotinamide (1-MNA) is detected as an abundant metabolite biomarker in nails of OSCC patients.

**Table 2:** Details on molecular docking by AutoDock Vina of Nicotine Imine upon selected methyltransferase enzymes involved in epigenetic-metabolic axis that support cancer growth and proliferation. Nicotine amine is detected as an abundant nicotine metabolized product in nails of OSCC patients.

| Sr. No | Name of Enzymes/Receptors and PDB ID | Binding Energy/Docking Score | RMSD Value l.b. | RMSD Value u.b. |
| --- | --- | --- | --- | --- |
| 1. | NNMT Enzyme (PDB ID: 3ROD) | -4.1 | 0.000 | 0.000 |
| 2 | Human Euchromatic Histone Methyltransferase 2 (PDB ID:2O8J) | -5.3 | 0.000 | 0.000 |
| 3. | Methyltransferase domain of human euchromatic histone methyltransferase 1 (PDB ID:4I51) | -4.7 | 0.000 | 0.000 |
| 4. | Catalytic domain of human DOT1L, a non-set domain nucleosomal histone methyltransferase (PDB id 1NW3) | -5.8 | 0.000 | 0.000 |
| 5. | Human euchromatic histone methyltransferase I SET Domain (PDB ID: 2IGQ) | -5.0 | 0.000 | 0.000 |
| 6. | Human DNMT1 (PDB id 4WXX) | **-6.2** | 0.000 | 0.000 |

**Table 3:** Details on molecular docking by AutoDock Vina of N-methylnicotinium ion upon selected methyltransferase enzymes involved in epigenetic-metabolic axis that support cancer growth and proliferation. N-methylnicotinium is detected as an abundant R-nicotine metabolized product in nails of OSCC patients.

| Sr. No | Name of Enzymes/Receptors and PDB ID | Binding Energy/Docking Score | RMSD Value l.b. | RMSD Value u.b. |
| --- | --- | --- | --- | --- |
| 1. | NNMT Enzyme (PDB ID: 3ROD) | -5.1 | 0.000 | 0.000 |
| 2 | Human Euchromatic Histone Methyltransferase 2 (PDB ID:2O8J) | -5.0 | 0.000 | 0.000 |
| 3. | Methyltransferase domain of human euchromatic histone methyltransferase 1 (PDB ID:4I51) | -4.9 | 0.000 | 0.000 |
| 4. | Catalytic domain of human DOT1L, a non-set domain nucleosomal histone methyltransferase (PDB id 1NW3) | -5.1 | 0.000 | 0.000 |
| 5. | Human euchromatic histone methyltransferase I SET Domain (PDB ID: 2IGQ) | -5.2 | 0.000 | 0.000 |
| 6. | Human DNMT1 (PDB id 4WXX) | **-5.4** | 0.000 | 0.000 |

**Figure 2.**
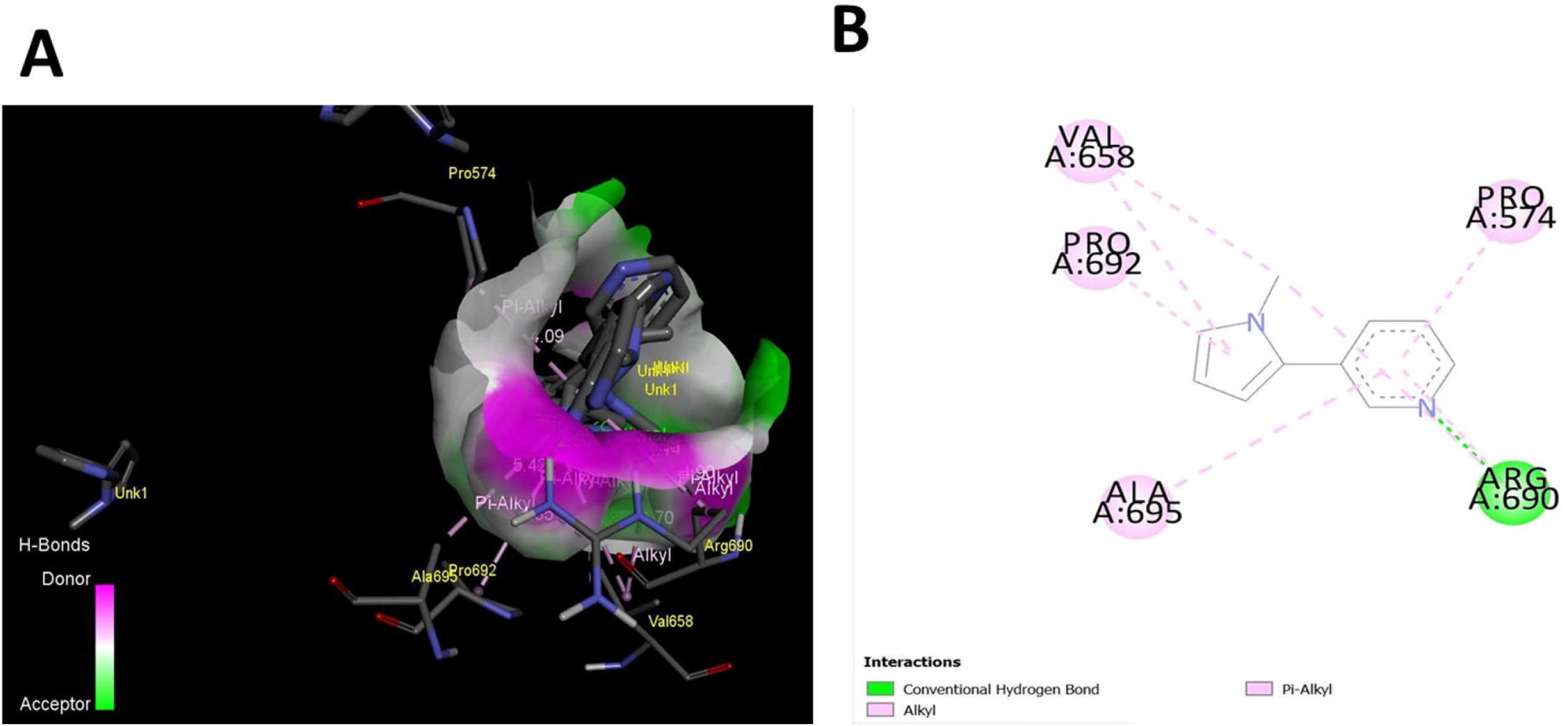
A nicotine derived metabolic product Nicotine imine (PubChem CID: 431) shows strong and specific binding to regulatory unit of DNMT1. **(A).** 3-D image generated by Discovery Studio Visualizer. **(B).** Ligand-Protein contacts generated as 2-D image by Discovery Studio Visualizer.

**Figure 3.**
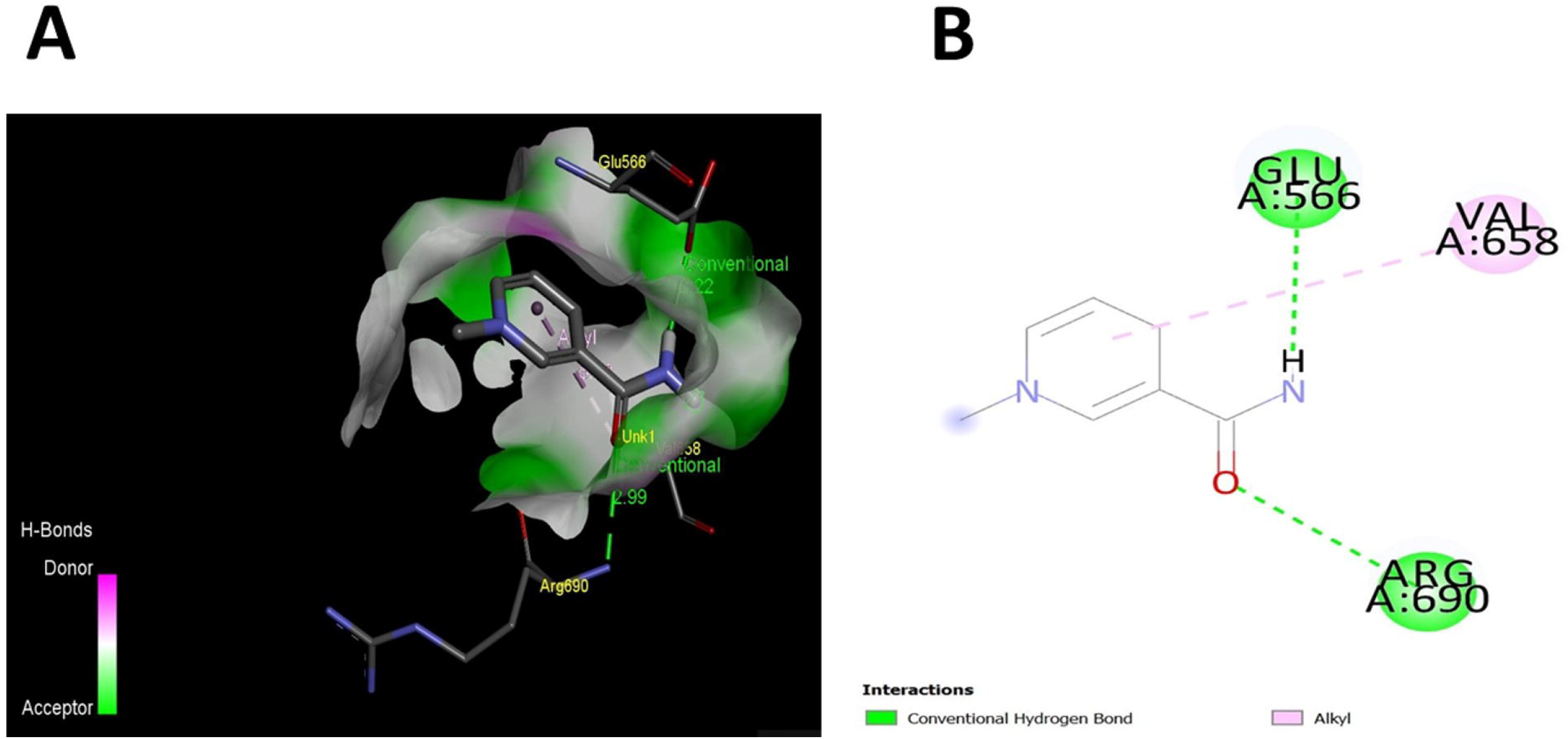
Oncometabolite 1-MNA generated by NNMT enzymes binds to regulatory domain of DNMT1. **(A).** 3-D image generated by Discovery Studio Visualizer**. (B).** Ligand-Protein contacts generated as 2-D image by Discovery Studio Visualizer.

**Figure 4.**
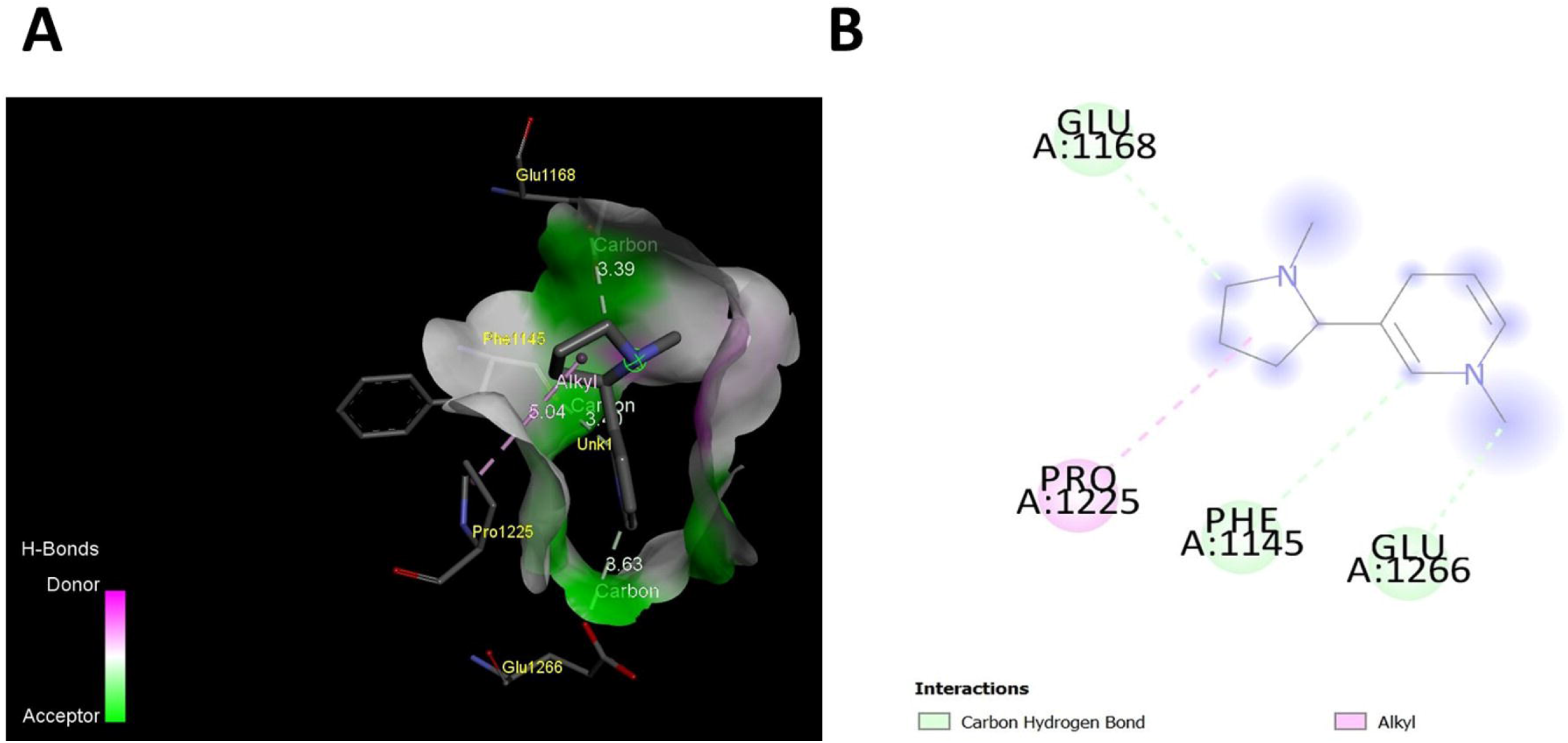
Oncometabolite N-methylnicotinium ion, a nicotine metabolized product to catalytic domain of DNMT1. **(A).** 3-D image generated by Discovery Studio Visualizer. **(B).** Ligand-Protein contacts generated as 2-D image by Discovery Studio Visualizer.

Furthermore, in case of a known DNMT1 inhibitor 5’-Aza-2’-deoxycytidine, specific interacting amino acid residues observed are ARG690, GLU572, SER570, PHE576 and PRO574 (Figure 5). Importantly, amino acid residues in case of DNMT1 inhibitor 5’-Aza-2’-deoxycytidine occupied the same regulatory domain as observed in case of nicotine imine and 1-MNA. Furthermore, similar nature of polar bonds and distance of polar bonds are recorded for nicotine imine, 1-MNA and DNMT1 inhibitor 5’-Aza-2’-deoxycytidine (Table 4).

**Table 4:** Determination of molecular interactions during molecular docking by AutoDock Vina and visualized by Discovery Studio Viewer.

| Name of Ligand | Binding<br>Energy/Docking<br>Score<br>(-Kcal/Mol) | Binding<br>positions (amino<br>acid residues) | Number of<br>Molecular<br>interactions | Distance<br>of polar<br>bonds |
| --- | --- | --- | --- | --- |
| 1-Methylnicotinamide<br>(PUBCHEM CID:<br>457) | 5.6 | 1. ARG-690<br>2. GLU-566<br>3. VAL658 | 1<br>1<br>1 | 2.99<br>2.22<br>3.5 |
| Nicotine imine<br>(PUBCHEM CID:<br>431) | 6.2 | 1. ARG690<br>2. PRO374<br>3. PRO692<br>4. VAL658<br>5. ALA695 | 1<br>1<br>1<br>1<br>1 | 2.7<br>2.6<br>4.99<br>4.10<br>3.5 |
| N-Methylnicotinium<br>ion<br>(PUBCHEM CID:<br>430) | 5.4 | 1. GLU1168<br>2. PHE-1145<br>3. PRO1225<br>4. GLU-1266 | 1<br>1<br>1<br>1 | 3.39<br>3.40<br>5.04<br>3.63 |
| 5'-Aza-2'-<br>deoxycytidine<br>(PUBCHEM CID:<br>22841634) | 7.0 | 1. ARG690<br>2. GLU572<br>3. SER570<br>4. PHE576<br>5. PRO574 | 1<br>1<br>1<br>1<br>1 | 2.2<br>2.5<br>2.9<br>3.5<br>3.5 |

**Figure 5.**
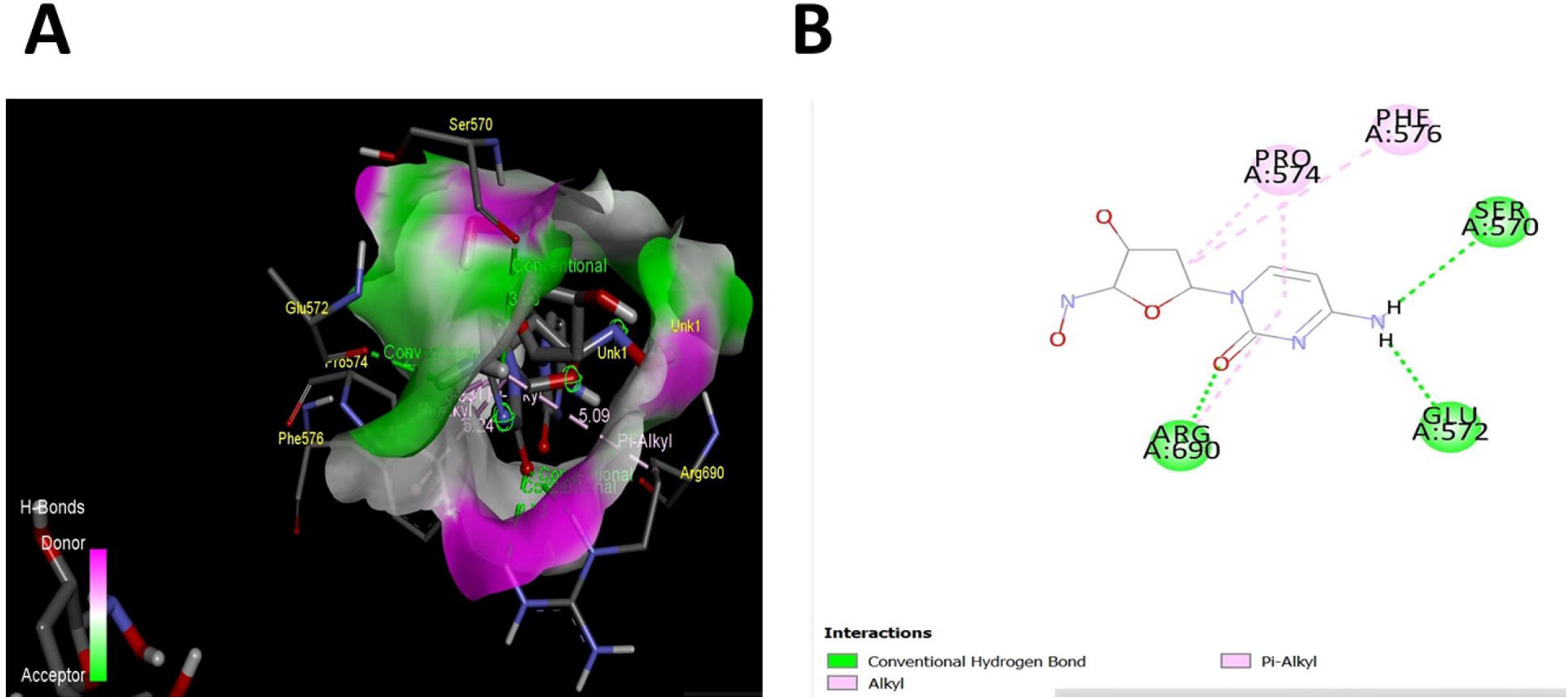
A known DNMT1 inhibitor 5’-Aza-2’-deoxycytidine (PUBCHEM CID: 22841634) shows specific binding to CXXC regulatory domain of DNMT1. **(A).** 3-D image generated by Discovery Studio Visualizer. **(B).** 5’-Aza-2’-deoxycytidine -DNMT1 contacts generated as 2-D image by Discovery Studio Visualizer.

Among the observed molecular docking data, we performed MD simulation for nicotine imine upon DNMT1. The collected data on RMSD plot of nicotine imine-DNMT1 (Figure 6A) and nicotine imine-DNMT1contacts (Figure 6B) is presented. The variation between RMSD plot of nicotine imine and DNMT1 is within the acceptable range of 1-3 Angstrom and that shows the efficient stability. Conversely, nicotine imine-DNMT1contacts show the involvement of similar amino acid residues including ARG-690, PRO574 as recorded in molecular docking and also in case of a DNMT1 inhibitor 5’-Aza-2’-deoxycytidine.

**Figure 6.**
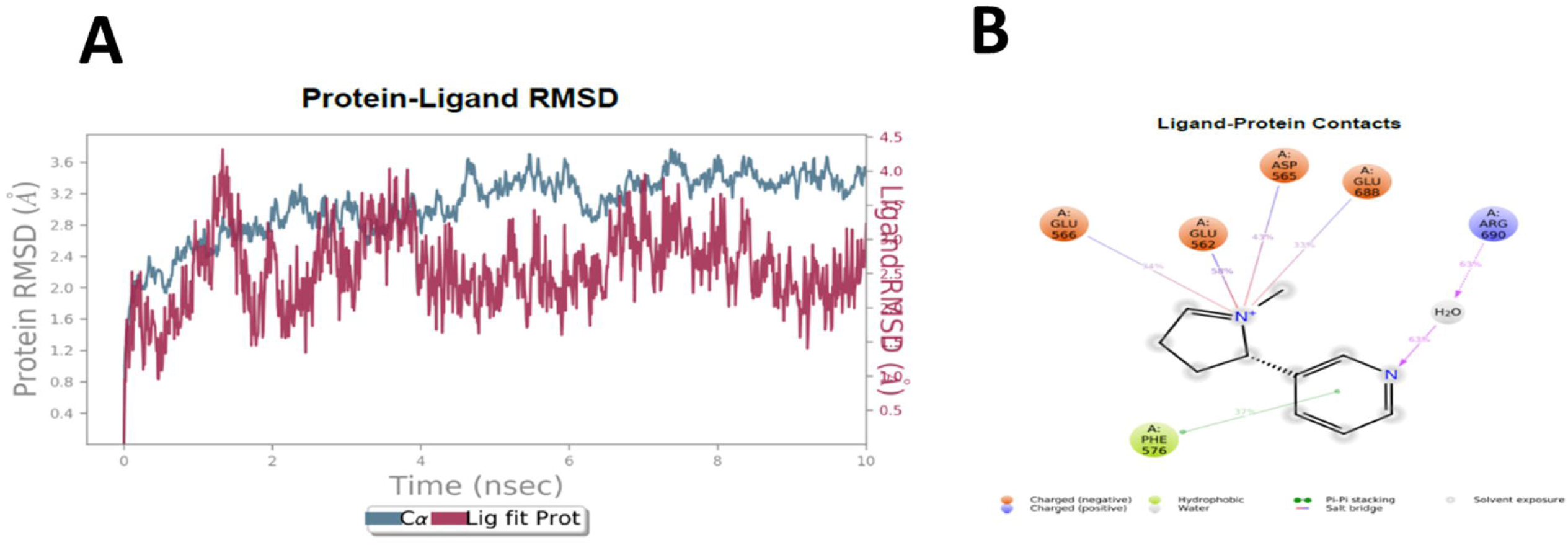
Protein-ligand Room Mean Square Deviation (RMSD) plot indicates the molecular interaction between Nicotine imine (PubChem CID: 431) and DNMT1 enzyme (PBD ID: 4WXX). **(A).** Here, left-Y axis represents the RMSD evolution of DNMT1 enzyme in view of structural conformation during simulation for duration 10 ns. Right-Y axis indicates the RMSD value of Nicotine imine (Lig-Fit-Pro) and this value is almost equivalent to the RMSD value of DNMT1. **(B).** DNMT1-Nicotine imine contacts profile.

## DISCUSSION

In recent appreciable data suggest on the contribution of metabolites generated during metabolic processes for their non-metabolic functions by translocating themselves into the nucleus. One of these functions is to regulate the epigenetic modifications and gene expression, which could in turn alter the metabolism in tumors and modify epigenetic scenario to contribute to malignant tumor growth (DeBerardinis et al., 2008; Hsu and Sabatini, 2008; Hanahan and Weinberg, 2011; Ulanovskaya et al., 2013; Feinberg et al., 2016; Yu et al., 2018; Kim et al., 2019).

In these cells, intracellular metabolic adaptations including activation of NNMT, methylation of nicotine are linked with the consumption of methyl group from SAM to methylate nicotinamide and nicotine derived products. Due to these changes, cancer cells achieve altered epigenetic state as global hypomethylation to promote expression of pro-tumorigenic genes (Feinberg and Vogelstein, 1983; Christmane et al.., 2002; Lopez-Serra and Esteller, 2008; Palii et al., 2008; Wu et al., 2008; Ulanovskaya et al., 2013; Kraus et al., 2014; Cui et al., 2020; Gissi et al., 2020). However, there is a significant gap on the suitability and availability of the nicotine metabolites and 1-MNA that influence the global DNA methylation and in turn pro-tumor microenvironment.

In the context of oncogenesis in OSCC, potential molecular pathways that converge together due to consumption of tobacco and gutkha products are not known. Our current understanding on the metabolized products of nicotine, a major constituent of chewing tobacco and tobacco related products in relation to OSCC is ambiguous. Furthermore, clinical detection of nicotine and nicotine-derived metabolites is limited in case of biological samples of various nature including non-conventional source like nails of OSCC.

## SAM, DNA METHYLATION and CANCER

Among plethora of detoxifying systems, there are reports on the overexpression of NNMT in OSCC and this cytoplasmic methyltransferase family of protein is known to consume SAM to produce 1-MNA from nicotinamide (Feinberg and Vogelstein, 1983; Christmane et al.., 2002; Lopez-Serra and Esteller, 2008; Palii et al., 2008; Wu et al., 2008; Ulanovskaya et al., 2013; Kraus et al., 2014; Cui et al., 2020; Gissi et al., 2020). Interestingly, the utilization of SAM by NNMT is suggested to lead to the DNA hypomethylation as a pro-tumor epigenetic landscape. But pertinent questions can be raised if 1-MNA, a metabolized product in OSCC is just a metabolic waste or it is further implicated in helping cancer cells to achieve favorable metabolic-epigenetic landscape. Simultaneously, the role of nicotine metabolized products such as nicotine imine and N-methylnicotinium ion in cancer cells as modulatory factors of metabolic-epigenetic axis that supports the growth and proliferation of OSCC is less studied.

Additionally, the use of SAM is indicated in the formation of N-methylnicotinium ion from the R-nicotine by an unknown class of methyltransferases in case of cancer cells (Nwosu et al., 1988; Berkman et al., 1995; von Weymarn et al., 2002; Kilgore et al., 2013; Jeltsch et al., 2016; Miletić et al., 2017; Yu et al., 2019). At the same time, S-nicotine is suggested to be metabolized into nicotine imine by the enzymatic action of cytochrome P450 family of enzymes (Nwosu et al., 1988; Berkman et al., 1995; von Weymarn et al., 2002; Kilgore et al., 2013; Jeltsch et al., 2016; Miletić et al., 2017; Yu et al., 2019). These data support the idea of existence of metabolic-epigenesis in cancer cells that supports growth and proliferation. However, detection of 1-MNA, nicotine imine and N-methylnicotinium ion at clinical levels is highly limited specifically in the nail of OSCC. The authors agree that reported novel approach on identification of nail metabolites also detects other routinely used drugs and metabolic products as shown in the TIC of healthy and OSCC subjects. Our lab and existing literature have attempted to address the accumulation of various types of drugs, metabolized drugs, environmental toxins (Oyoo-Okoth et al., 2010; Krumbiegel et al., 2016; Mitruka et al., 2020; Sharma et al., 2020). However, this finding presents a first report on the detection of oncometabolites such as 1-MNA, nicotine imine and N-methylnicotinium ion and their possible link with the metabolic-epigenetic signaling axis. In this paper, we show the abundance of 1-MNA, nicotine imine and N-methylnicotinium ion in nails of OSCC compared to healthy individuals. In essence, our data strengthen the basis of cytoplasmic conversion of nicotinamide and nicotine into intracellular oncometabolites by using SAM. Due to its high use in the cytoplasmic enzymatic conversion, SAM based DNA methylation by DNMT1 is compromised and that leads to global hypomethylation.

Cellular relevance of these oncometabolites namely 1-MNA, nicotine imine and N-methylnicotinium ion is missing and these may be connected with the protumoral metabolic-epigenomic axis. We report here on the in-silico approach to link the molecular interactions of these oncometabolites with various methyltransferase enzymes including NNM1, DNMT1 and histone methyltransferase.

## IN SILICO EVIDENCE AND DNMT1 AND NICOTINE PRODUCTS

Upon performing molecular docking and visualization using Discovery Studio Viewer, it is revealed that nicotine imine interacts with the key amino acid residues namely ARG690, PRO574, VAL658, PRO692 and ALA695 of DNMT1. Existing data support that various known DNMT1 inhibitors including 5’-Aza-2’-deoxycytidine also bind to the similar regulatory domain spanning from 550 to 700 amino acid residues that actually influence the methylation activity of DNMT1 (Feinberg and Vogelstein, 1983; Christmane et al.., 2002; Lopez-Serra and Esteller, 2008; Palii et al., 2008; Gissi et al., 2020). A comparison of molecular interactions and similar amino acid residues between nicotine imine and 5’-Aza-2’-deoxycytidine clearly convinces the possibility that nicotine imine is capable of inhibiting the activity of DNMT1. The binding affinity of nicotine imine is comparable to the known DNMT1 inhibitor 5’-Aza-2’-deoxycytidine. Besides nicotine imine, 1-MNA that is detected as an oncometabolite in the nail of OSCC is also able to interact specifically at the same regulatory domain as in the case of nicotine imine and 5’-Aza-2’-deoxycytidine.

In this investigation, we have used *in silico* approach to understand the molecular interactions of 1-MNA, nicotine imine and N-methylnicotinium ion with DNMT1 that plays a key role in the global DNA methylation. Concurrently, DNA hypomethylation is known as a key pro-tumor epigenetic state in cancer cells including OSCC. The molecular simulation dynamics data by RMSD evolution plot of DNMT1 and nicotine imine shows the fluctuations in the RMSD values within the order of 1-3Å and that is within the limit for the stability of target proteins bound by ligand. Taken together, molecular docking, simulation and dynamics studies support the role of nicotine imine as an inhibitor of DNMT1 by binding to the regulatory domain.

DNMT1 is known to maintain the DNA methylated epigenetic state in both cancer and normal cells (Feinberg and Vogelstein, 1983; Christmane et al.., 2002; Lopez-Serra and Esteller, 2008; Palii et al., 2008; Gissi et al., 2020). Furthermore, DNMT1 contains two major components as c-terminus catalytic and N-terminus regulatory domains. Within the regulatory N-terminus domain, CXXC domain (550-700 amino acid residues) is known for the allosteric regulation of DNMT1 catalytic activity. There are appreciable reports on the development of pharmacological inhibitors including 5’-Aza-2’-deoxycytidine that is known to bind to the same CXXC domain and act as an inhibitor of DNMT1 (Feinberg and Vogelstein, 1983; Christmane et al.., 2002; Lopez-Serra and Esteller, 2008; Palii et al., 2008; Gissi et al., 2020).

Our molecular docking data provide additional insights on the generated N-methylnicotinium ion, a product of R-nicotine as an inhibitor of the catalytic site of DNMT1. Furthermore, these data indicate that catalytic site amino acid residues including GLU1168, PHE-1145, PRO1225 and GLU-1266 are bound by N-methylnicotinium ion. There is in silico and in vitro data on the use of GLU1168, PHE-1145, PRO1225 and GLU-1266 as important enzymatic residues by DNMT1 and inhibitors are reported that bind to the similar target sites (Feinberg and Vogelstein, 1983; Christmane et al.., 2002; Lopez-Serra and Esteller, 2008; Palii et al., 2008; Gissi et al., 2020). In fact, this is the first proposition on the molecular basis of R-nicotine metabolized product N-methylnicotinium ion which can bind to the catalytic site of DNMT1 and induce DNA hypomethylation in cancer cells including OSCC.

Our data also suggest that abundance of oncometabolites 1-MNA, nicotine imine and N-methylnicotinium ion in OSCC helps cancer cells in two possible pro-tumor pathways. One is in the form of less availability of SAM for DNMT1 to carry out DNA methylation that leads to reduced global DNA methylation and creates pro-tumor epigenetic events. There are reports that suggest the connection between global DNA hypomethylation and pro-tumor events that support cancer cells. Hence, our work accentuates the similar understanding with new evidence on the role of these oncometabolites in relation to DNA hypomethylation. Second molecular pathway by these oncometabolites 1-MNA, nicotine imine and N-methylnicotinium ion that we propose is that DNMT1 enzyme activity is inhibited due to the molecular interaction at regulatory and catalytic domains as predicted by molecular docking and MD Simultion study.

Based on the molecular docking and MD Simulation study, 1-MNA and nicotine imine are predicted to bind to the regulatory domain of DNMT1 compared to other methyltransferase enzymes. At the same time, N-methylnicotinium ion is shown to bind with the catalytic site of DNMT1. Our data thus, is in consonance with the reported molecular interaction studies that showed the similar binding sites by 5’-Aza-2’-deoxycytidine a known anticancer compound and inhibitor of DNMT1 (Feinberg and Vogelstein, 1983; Christmane et al.., 2002; Lopez-Serra and Esteller, 2008; Palii et al., 2008; Gissi et al., 2020).

Altogether, our molecular docking and MD Simultion data encourage us to predict that generation of nicotine imine and 1-MNA in cancer cells is an efficient metabolic adaptation to meet the growth and proliferation by employing two distinct pathways. One possibility is that the SAM, a key methyl-donor that is consumed during the nicotine metabolism and generation of 1-MNA may help cancer cells to gain pro-tumor DNA hypomethylation. On the other hand, accumulation of nicotine metabolized products including nicotine imine and N-methylnicotinium ion along with 1-MNA pose a blockade for DNMT1 during the DNA methylation process. In both ways, the required epigenetic state is created within the cancer cells as global DNA hypomethylation and in turn it favors the cancer cells for better growth and proliferation. A proposed model on the implications of these oncometabolites including 1-MNA, nicotine imine and N-methylnicotinium ion is presented in Figure 7.

**Figure 7.**
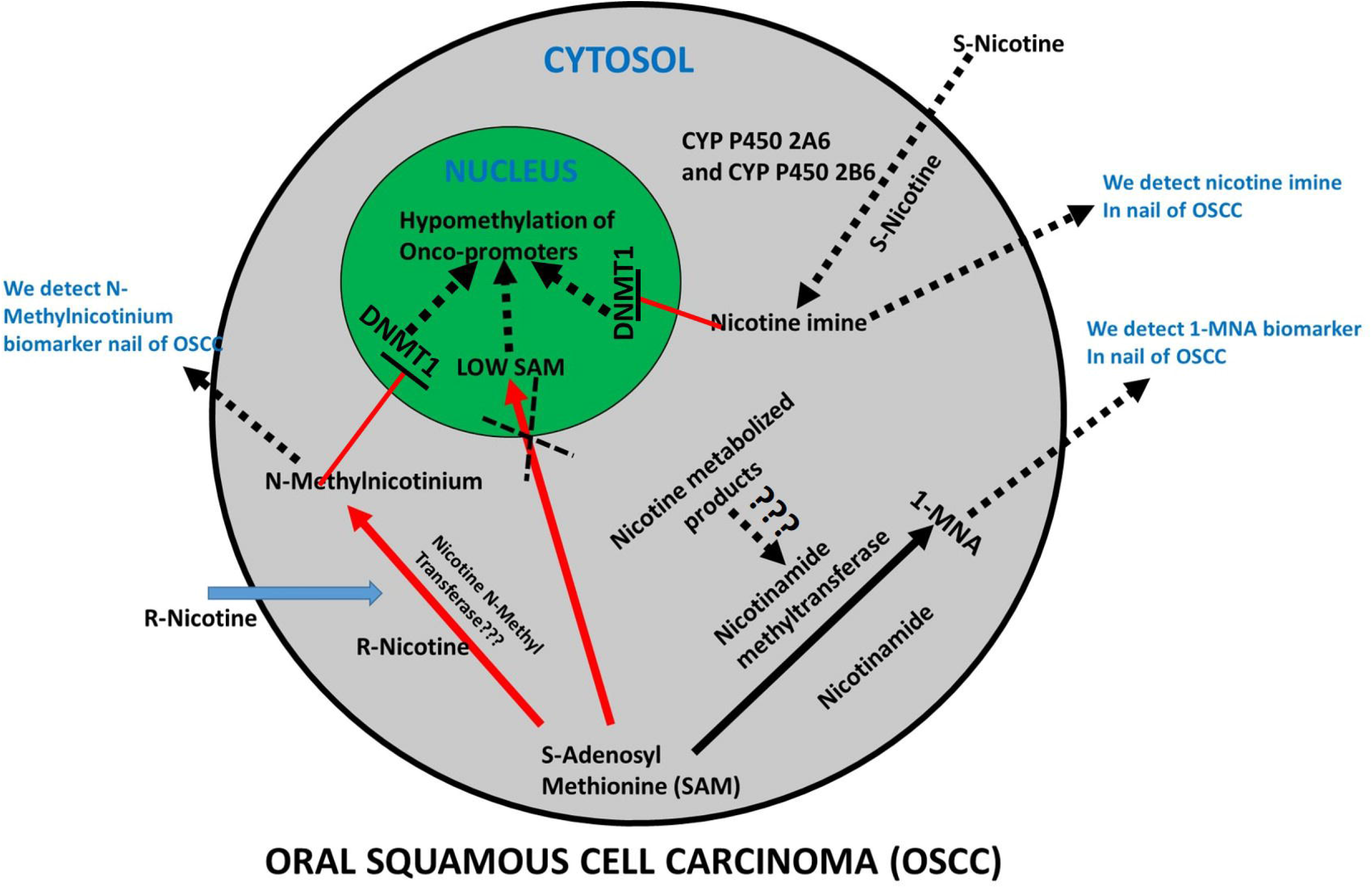
A proposed model on the role of nicotine derived metabolites in the methylation process during epigenetic changes which occur in cancer cells. These metabolites promote metabolic reprogramming by modulating epigenetic enzyme DNMT1.

## LIMITATIONS AND FUTURE SCOPE

The present study is performed on limited sample size due to constraints of the financial resources. The selection of subjects in both healthy and OSCC groups ensured the proportionate representation of similar age and gender distribution. The authors agree that sub-grouping of OSCC patients was not done based on the different grades and the same is not highlighted in this paper. However, this does not jeopardize establishment of the contention obtained in the present study. For further strengthening of the proposed hypothesis, future studies on larger sample size with inclusion of premalignant lesions and subgrouping of OSCC based on their grades are warranted. Although the present study provided a strong proof of concept for establishment of predictive and diagnostic biomarker, an appropriate disease progression model that reflects the true continuum of malignant transformation is needed. Here, it is important to highlight that VTGE approach may be employed to study intracellular metabolized nicotine products and other metabolic adaptations in cell line models and that may reveal the additional information in oral cancer to link nicotine, metabolic reprogramming and epigenetic states. In future, cellular, preclinical and clinical studies are warranted that may address the role of nicotine metabolized products in driving cellular and microenvironmental adaptations that support tumorigenesis.

## CONCLUSION

The presence of N-methylnicotinium ion, nicotine imine and 1-MNA identification in nail samples indicates their potential as predictive and detectable biomarkers for OSCC. Additionally, 1-MNA a metabolic product due to high activity of NNMT and use of SAM is proposed as potential oncometabolites detected in nails of OSCC. In fact, a strong metabolic clue within OSCC is also proposed that is, generation of these metabolites including nicotine imine, N-methylnicotinium ion and 1-MNA creates a deficiency/lack of SAM for DNA methylation that is linked to the global DNA methylation. Bsed on various in silico analyses, we propose that these metabolic products including nicotine imine, N-methylnicotinium ion and 1-MNA, besides being potential metabolite biomarkers in nails of OSCC, help cancer cells to achieve global DNA hypomethylation states by blocking the activity of DNMT1, rather than them being simply metabolic wastes.

## Data Availability

Data will be made available.

## FUNDING SOURCE

The authors acknowledge financial support from Dr. D.Y. Patil Vidyapeeth, Pune, India (DPU/05/01/2016).

## ETHICAL STATEMENT

In this study, Institutional Ethics Committee of Dr. D. Y. Patil Vidyapeeth, Pune (Ref.NoDYPV/EEEEEC/245/2019) approval was obtained.

## Details of figures and their legends

**Figure S1.**
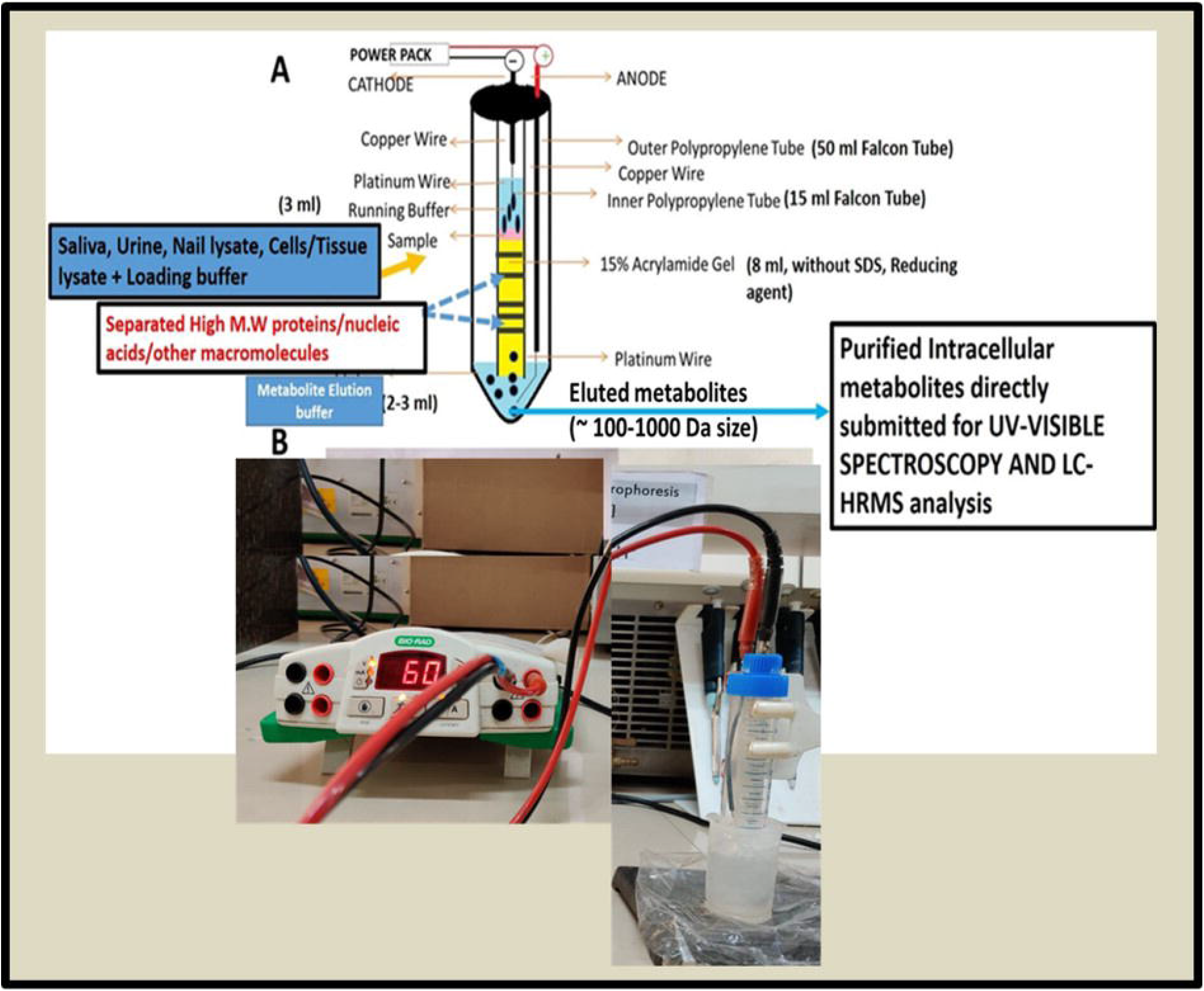
A flow diagram of novel and specifically designed vertical tube gel electrophoresis (VTGE) system for intracellular metabolite purification. **(A)** An assembly and design of VTGE system is illustrated. **(B)** A working model is depicted for VTGE system.

**Figure S2.**
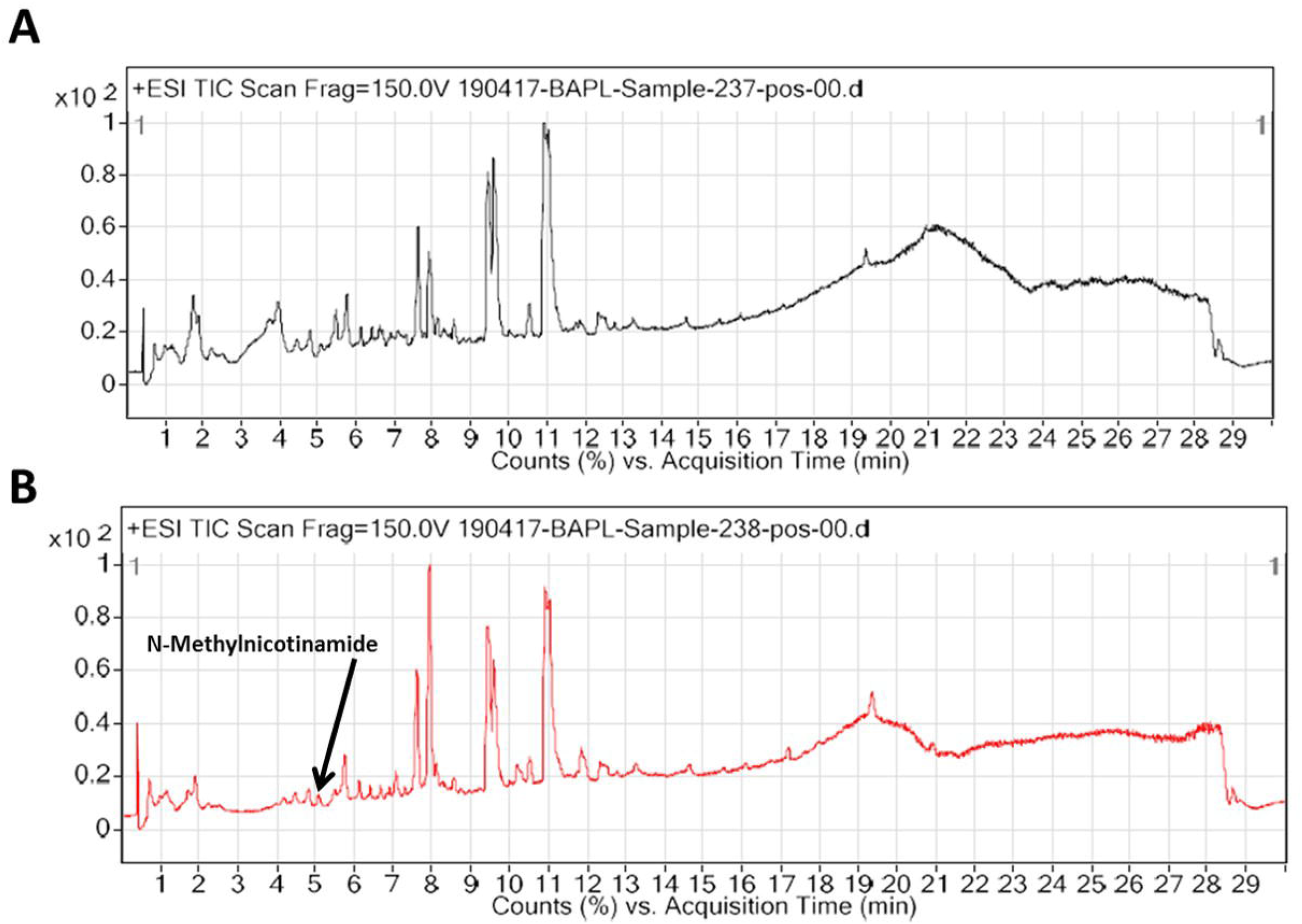
VTGE purified nail metabolites identified by LC-HRMS show distinct water loss adduct TIC in OSCC over healthy subjects with the presence of oncometabolite N-methylnicotinamide. For the preparation of TIC of nail metabolites, purified nail metabolites of healthy and OSCC subjects were submitted to LC-HRMS. **(A).** A representative water loss adduct TIC of Healthy subject is presented in positive ESI mode Water loss adduct chromatogram. **(B).** A representative sodium adduct TIC of OSCC is presented in positive ESI mode and shows the presence of oncometabolite N-methylnicotinamide.

**Figure S3.**
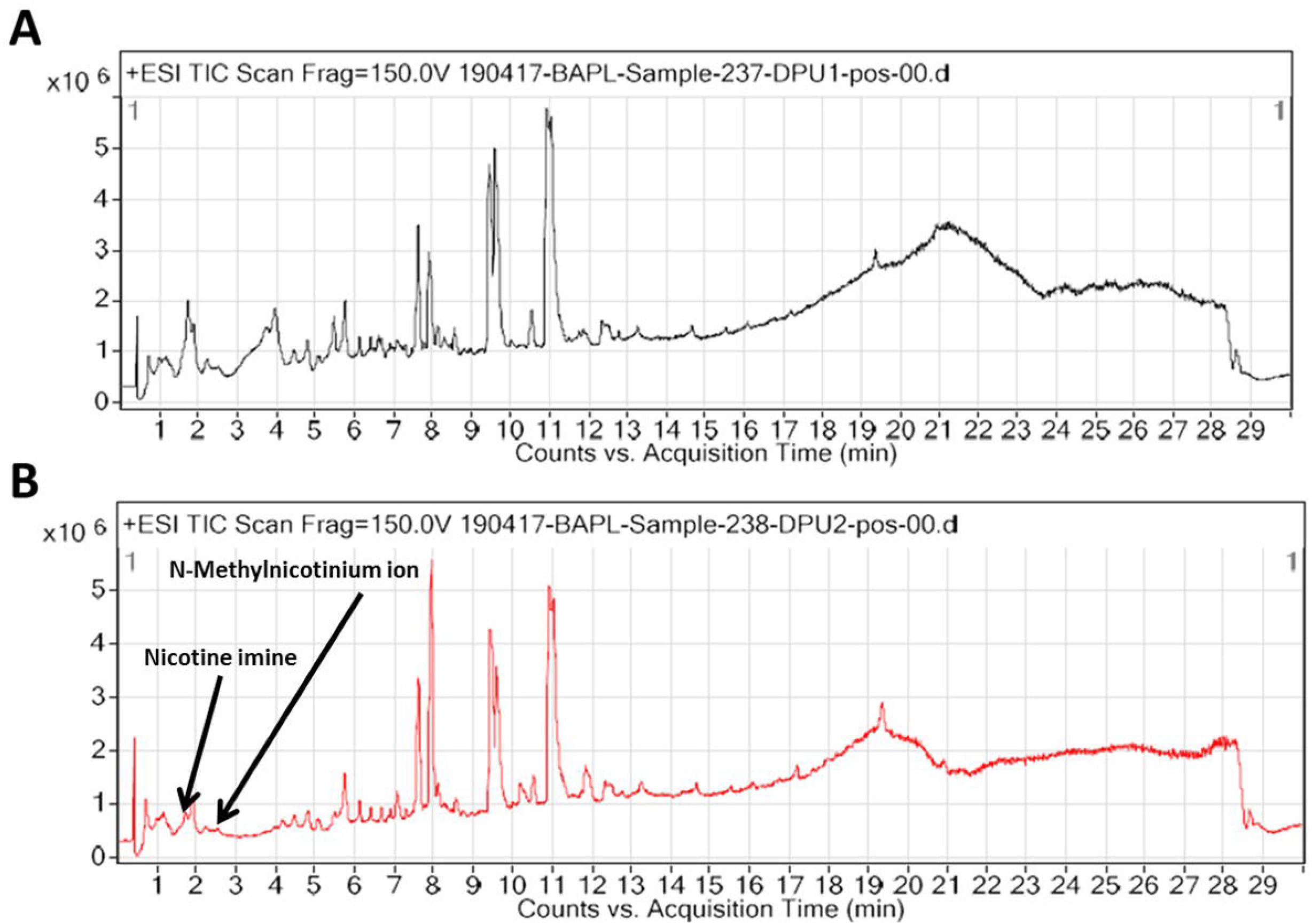
VTGE purified nail metabolites identified by LC-HRMS show distinct Sodium adduct TIC in OSCC over healthy subjects with the presence of Nicotine Imine and N-Methylnicotinium ion. For the preparation of TIC of nail metabolites, purified nail metabolites of healthy and OSCC subjects were submitted to LC-HRMS. **(A)** A representative water loss adduct TIC of Healthy subject is presented in positive ESI mode. **(B)** A representative sodium adduct TIC of OSCC is presented in positive ESI mode and shows the presence of nicotine metabolized product Nicotine Imine and N-Methylnicotinium ion.

## Notes

### Competing Interest Statement

The authors have declared no competing interest.

### Author Declarations

Institutional Ethics Committee of Dr. D. Y. Patil Vidyapeeth, Pune (Ref.NoDYPV/EEEEEC/245/2019) approval was obtained before commencement of the study.

